# Impact of low-dose intrathecal morphine on orthopedic surgery: a protocol of a systematic review and meta-analysis of randomized controlled trials

**DOI:** 10.1101/2022.02.12.22270830

**Authors:** Yue Lei, Mu Guanzhang, Lin Zengmao, Sun Haolin

## Abstract

**Introduction:** Patients undergoing orthopedic surgery usually suffer considerably from peri-operative pain and intrathecal morphine (ITM) has recently been used as an effective analgesic method. The intrathecal morphine dose achieving optimal analgesia for orthopedic surgery while minimizing side effects has not yet been determined. There is currently a lack of literature synthesis on the safety and effects of low-dose ITM on orthopedic surgery.

**Methods and analysis:** A thorough literature search will be performed on multiple electronic databases and trial registries until January 11^th^ 2022, and reference lists will be examined. Two independent reviewers will select studies, extract data and assess the risk of bias and quality of the included studies. We will select randomized controlled trials comparing low-dose ITM (≤100μg) with high-dose ITM (≥200μg) or placebo treatment or other comparatives. We will assess the risk of bias using the Cochrane risk of bias tool and Jadad score will be applied to assess the quality of each included trial. We will also use Grading of Recommendations, Assessment, Development, and Evaluation (GRADE) to determine the confidence in effect estimates. Publication bias will be evaluated by visually inspecting funnel plots and Egger regression asymmetry test in estimations of more than 10 trials. Standard meta-analysis will be performed using R package meta.

**Ethics and dissemination:** The results of this systematic review and meta-analysis will be disseminated through peer-reviewed journal and related conferences. The data used in this meta-analysis will not contain individual patient data and ethical approval is therefore not required.

## INTRODUCTION

Postoperative pain management is a significant public health concern globally, especially for orthopedic surgery. In clinical practice, patients undergoing orthopedic surgery usually suffer considerably from pain that exists preoperatively, intraoperatively, and postoperatively. Besides, pain can appear as a postoperative complication and may become chronic and last for a long time. Hence, post-surgical analgesia is critical for restoring mobility and maintaining patient satisfaction in the early postoperative period. Traditional peri-operative pain management, effective as it is, relies heavily on opioids orally or intravenously, which may cause overdose of opioids and subsequent side-effects or opioid dependence[1]. Managing pain efficiently with minimal morphine consumption is the goal that orthopedic surgeons and anesthetist should strive to achieve.[2]

Intrathecal morphine (ITM) has been proven to be an ideal solution, firstly induced to clinical application in 1979, as the drug is administered directly into the cerebrospinal fluid, where the opioid acts close to the central nervous system, is effectively performed at dosage of typically no more than 1 mg of morphine [3, 4]. However, previous evidence has shown that both efficacy and side-effects are dose-dependent in various surgical scenarios[4-6]. Therefore, weighing the benefits and potential side effects should be a primary concern when determining the dosage of ITM in clinical use. Compared with a higher dosage, administration of low-dose morphine (≤100 μg) intrathecally has been validated as the optimal dosage by previous studies, for its effectiveness with minimal side-effects [6, 7]. In recent decades, multiple randomized controlled trials have emerged, proving the efficacy and safety of systematic use of low-dose ITM on patients undergoing orthopedic surgeries, and therefore an updated synthesis of the literature is needed.

## OBJECTIVE

To date, no similar systematic review was found as the International Prospective Registry of Systematic Reviews (PROSPERO) and Cochrane Database of Systematic Reviews. This systematic review aims to identify and critically evaluate randomized controlled trials of systematic use of low-dose ITM (≤100 μg) for patients undergoing orthopedic surgery. A comprehensive understanding of the current level of evidence in the literature will help clarify the clinical utility of low-dose ITM on orthopedic surgery and inform future research.

## METHODS AND ANALYSIS

The protocol was developed according to the Preferred Reporting Items for Systematic Review and Meta-Analysis Protocols (PRISMA-P) and the Cochrane Handbook for Systematic Reviews of Interventions [8, 9].

### Characteristics of the studies

The literature search will include studies according to the review’s consensus-based criteria, objectives and clinical questions involving four aspects: participants, interventions, comparisons, and outcomes (PICO). RCTs that compared low-dose ITM (≤100 μg of morphine) versus high-dose ITM (≥200μg of morphine) or other non-morphine treatment on orthopedic surgery will be selected according to the recommendations of the Cochrane Group. No language limit will be applied. Non-RCT studies, such as non-randomized cohort studies, case-control studies, case series, case reports, narrative reviews, editorials, or animal researches, will not be included.

### Eligibility criteria

Studies will be selected for inclusion if their participants meet specific criteria:

- Patients who underwent orthopedic surgeries, including spinal surgery, joint surgery, bone fracture surgery, or surgery for bone tumors;
- Low-dose ITM was implemented (≤100 μg of morphine).

We will exclude studies that included participants with:

- Comparison group that used morphine administered via other approaches (e.g., intravenous, subcutaneous, or oral) or comparison group of unclear contrast;
- Participants who underwent non-orthopedic surgeries.

### Outcome measures

#### Major outcomes

- The cumulative dose of analgesics at 24 h postoperatively (converted to morphine equivalent according to Opioid Equivalence Chart by NHS[10]).
- Incidence of common opioid-related side-effect

#### Minor outcomes

- Pain intensity at the first 12-24 h after surgery.
- Time to first analgesic requirement after the operation.
- The proportion of patients required rescue analgesics post-operatively.
- Intraoperative blood loss.

### Patient and public involvement

This research will be conducted without patient involvement. Patients will not be invited to participate in study design, results interpretation, or the writing process of the current review.

### Search strategy

We will search the following electronic databases, registries and websites on January 11^th^ 2022, unrestricted by date. Grey literature and non-English studies will be not excluded.

- *English Databases:* PubMed, Cochrane library and Web of science,
- *Chinese database:* Cnki.net
- *Trial registries:* ClinicalTrials.gov.

The reference lists of retrieved trials and previous systematic reviews will be searched for citations of potentially eligible trials. In case any questions about trials arise, the corresponding author of the articles will be contacted. The search strategy is shown in Table.1.

**Table 1.** Search terms of retrieval of studies

|  | Search terms |
| --- | --- |
| PubMed | ((“orthopedic” or “hip” or “knee” or “shoulder” or “elbow” or “ankle” or “wrist” or “extremities” or “spinal” or “intervertebral” or “lumbar” or “cervical” or “scoliosis”) and (“operation” or “instrumentation” or “reduction” or “fixation” or “fusion” or “surgery” or “replacement” or “discectomy” or “arthroplasty” or “implant” or “prosthesis” or “osteotomy” or “arthroscopy”)) and (“intrathecal morphine” or “intrathecal opioids”) |
| Cochrane library | #1: (“orthopedic” OR “hip” OR “knee” OR “shoulder” OR “elbow” OR “ankle” OR “wrist” OR “extremities” OR “spinal” OR “intervertebral” OR “lumbar” OR “cervical” OR “scoliosis”): ti, ab, kw<br>#2: (“operation” OR “instrumentation” OR “reduction” OR “fixation” OR “fusion” OR “discectomy” OR “surgery” OR “replacement” OR “arthroplasty” OR “implant” OR “prosthesis” OR “osteotomy” OR “arthroscopy”): ti, ab, kw<br>#3: (“intrathecal morphine” OR “intrathecal opioids”): ti, ab, kw<br>#4: #1 AND #2 AND #3 |
| Web of Science | TS=((“orthopedic” or “hip” or “knee” or “shoulder” or “elbow” or |
|  | “ankle” or “wrist” or “extremities” or “spinal” or “intervertebral” or “lumbar” or “cervical” or “scoliosis”) and (“operation” or “instrumentation” or “reduction” or “fixation” or “fusion” or “surgery” or “replacement” or “discectomy” or “arthroplasty” or “implant” or “prosthesis” or “osteotomy” or “arthroscopy”)) AND TS=(“intrathecal morphine” or “intrathecal opioids”) |
| ClinicalTrials.gov | All studies; Condition or disease: orthopedic disorders; Other terms: intrathecal morphine |

\*The search strategy for Chinese literature is provided in Supplementary file 1.

### Selection of studies

Two independent reviewers (Y.L. and M.G.Z.) will screen the titles and abstracts of the enrolled studies and irrelevant studies will be excluded. Trials selected by the first selection will then be read in full-length article for a second selection. Authors will be contacted if clarification about study design is required. Any occurring disagreements will be noted and then discussed by the whole research group to reach a consensus. The selection process will be presented in a PRISMA flow diagram (Fig. 1) [11].

### Data extraction and management

Two independent reviewers (Y.L. and M.G.Z.) will extract data in accordance with the Cochrane Collaboration data extraction form. An independent double-check process will be undertaken by a third reviewer (L.Z.M or S.H.L.) when the extraction process has been finished.

### Assessing the methodological quality

The risk of bias for each included RCTs will be assessed by two reviewers (Y.L. and M.G.Z.) independently using the Cochrane risk of bias tool, and the overall quality of each included trial will be assessed by Jaded score[12]. Any disagreement will be resolved by the consensus of the whole group. The graphical presentation of the assessment of the risk of bias will be generated by RevMan 5.3.

We will also apply the Grading of Recommendations, Assessment, Development, and Evaluation (GRADE) approach to evaluate the overall quality of the evidence-based on five domains: limitations of design, inconsistency of results, indirectness, imprecision, and other factors (e.g., publication bias). GRADE approach evaluates the quality of evidence as ‘high’, ‘moderate’, ‘low’, or ‘very low’ by the outcomes [13].

### Dealing with missing data

For articles that did not describe the sample size or results as a mean and SD or standard error of the mean and 95% confidence interval, we will contact the corresponding author for relevant raw data via email. If the corresponding author failed to reply, we will take the median (IQR) as approximations of the mean (SD), by estimating the mean as equivalent to the median, and the SD as the IQR divided by 1.35, or the range divided by 4 as Gonver et, al. did [6].

### Data synthesis and analysis

The results from finally screened studies will be combined to estimate as effective results in standardized mean differences (SMD) and 95% CI for continuous outcomes. As to dichotomous outcomes, pooled risk ratio (RR) and 95% CI will be estimated. The synthesis will be done by generating a forest plot of the study estimates. We will evaluate the heterogeneity of the included studies with I^2^ test. Heterogeneity will be examined by I^2^ value as low, moderate or high (I^2^ value of 25%, 50% and 75% respectively). Statistical significance will be set at P<0.05 in this review.

### Assessment of publication biases

If there are over 10 trials included in the meta-analysis, reporting bias will be examined by constructing funnel plots and performing Egger regression asymmetry test[14].

### Sensitivity analysis

To confirm the robustness of our findings, a sensitivity analysis will be conducted by omitting the data from the trial one by one from the pooled analysis.

## Data Availability

All data produced in the present work are contained in the manuscript

## AUTHOR CONTRIBUTIONS

Y.L. concepted the research questions, designed the search strategy and prepared the manuscript draft. M.G.Z. designed search strategy, edited the manuscript. Y.L. and M.G.Z. will independently screen the potential studies, extract data and assess the risk of bias from included studies. Y.L. and M.G.Z. revised the search strategy and the manuscript. L.Z.M. and

S.H.L. revised the manuscript and approved the final version and will arbitrate potential disagreements. All authors approved the final version of this manuscript.

### FUNDING

This study is granted by Interdisciplinary clinical research project of Peking University First Hospital (Grant number: 2021CR31).

### PATIENT CONSENT

Not required.

### ETHICS APPROVAL

Not required.

### DATA SHARING STATEMENT

Data are available upon reasonable request.

